# Cardiometabolic disorder comorbidity within affective disorders: analysis of 1.6 million participants in the Our Future Health cohort

**DOI:** 10.1101/2025.11.23.25340839

**Authors:** Duncan Swiffen, Arish Mudra Rakshasa-Loots, Amber Roguski, Christina Steyn, Katie F. M. Marwick, Kelly Fleetwood, Daniel J Smith

**Author notes:** Corresponding author (DS).

## Abstract

**Background:** Affective disorders - including bipolar disorder, depression and anxiety disorders - are associated with increased risk of cardiometabolic disorders and premature mortality.

**Aim:** To assess prevalence and the association of cardiometabolic comorbidity in people with bipolar disorder, major depression and anxiety disorders within the Our Future Health population cohort.

**Method:** We analysed associations between affective and cardiometabolic disorders in 1,584,273 UK-based adults. Participants were split into three affective disorder groups based on self-reported lifetime diagnoses: bipolar disorder (n = 8,555); depressive disorders (n = 275,746); and anxiety disorders (n = 49,645); plus a comparison group of participants with no self-reported history of mental health problems (n = 1,250,327). For each group, we calculated age-sex standardised prevalence and sex-stratified standardised prevalence of self-reported lifetime diagnoses of any cardiometabolic disorder, nine individual cardiometabolic disorders and cardiometabolic multimorbidity. We calculated odds ratios for cardiometabolic disorders for each group, adjusted for age, sex, ethnicity, household income, smoking status, alcohol consumption, activity levels, chronotype and educational attainment.

**Results:** The standardised prevalence [95% confidence interval] of any cardiometabolic disorder within each of the affective disorders groups was higher than in the comparison group. For bipolar disorder this was 41.14% [39.36, 42.93], for depressive disorders 36.75% [36.49, 37.02] and for anxiety disorders 28.79% [28.38, 29.38], compared to 24.75% [24.66, 24.83] in the comparison group. There were sex-specific differences, with the prevalence of all cardiometabolic disorders except obesity elevated more in males than in females across each affective disorder group. Adjusted odds ratios of each of the nine individual cardiometabolic disorders were increased for bipolar disorder and for depressive disorders, and the adjusted odds of all except myocardial infarction were increased for anxiety disorders, albeit at a lower magnitude. For example, for type II diabetes, the adjusted odds ratios [95% confidence intervals] were 2.75 [2.46, 3.08] in bipolar disorder, 1.99 [1.93, 2.05] in depressive disorders and 1.11 [1.02, 1.21] in anxiety disorders, and for hypertension this was 1.62 [1.50, 1.76] in bipolar disorder, 1.62 [1.59, 1.65] in depressive disorders and 1.31 [1.27, 1.36] in anxiety disorders.

**Conclusions:** We identified high rates of cardiometabolic comorbidity for bipolar disorder, depression and anxiety disorders within a cohort of 1.6 million participants. These associations were independent of a range of potential confounding factors. Future work should seek to understand why risk of cardiometabolic disorder is particularly high in people with bipolar disorder and in men.

## Introduction

Affective disorders - including bipolar disorder, depression and anxiety disorders - are associated with significant global burden of disease (1) and premature mortality (2). Cardiometabolic disorders such as angina, coronary artery disease, myocardial infarction, congestive heart failure, stroke, type 2 diabetes, hypertension, hypercholesterolaemia and obesity are common in the general population (3), but are seen with increased prevalence in people with affective disorders (4–6) and are associated with poorer mental and physical health outcomes (7, 8). Evidence from genetic (9, 10) and longitudinal (11, 12) studies suggests that the relationship between affective and cardiometabolic disorders is likely to be bidirectional and may involve overlapping pathophysiological mechanisms (13).

Large cohort studies allow for complex associations and putative transdiagnostic mechanisms to be explored. Our Future Health (OFH) is the largest ever population health research programme that, as of May 2025, includes almost 1.8 million participants aged 18 years and over (9). A specific ambition of OFH is to recruit a sample that is reflective of the UK population in terms of age, sex, ethnicity and index of multiple deprivation (10). Our goal was to make use of data from the OFH cohort to assess in detail the relationships between affective disorders and cardiometabolic disorders, with a view to replicating previous work and generating new hypotheses on potential mechanisms of comorbidity.

## Materials and methods

### Participants

Participants were aged 18 years or older and living in the UK. Recruitment to OFH is ongoing and started in July 2022 through the UK National Health Service (NHS) DigiTrials service ([Recruitment Service - NHS England Digital](https://digital.nhs.uk/services/nhs-digitrials/recruitment-service)), community pharmacy networks, the NHS Blood and Transplant service, and pre-existing NHS primary and secondary care programmes (e.g. the primary care-based Health Check Programme) (10). Individuals were asked to volunteer mainly through postal invitations or via the OFH website (www.ourfuturehealth.org.uk).

All participants provided informed consent through completion of an electronic informed consent form prior to taking part in the study. All study procedures involving human participants received ethical approval from the Cambridge East NHS Research Ethics Committee (ref: 21/EE/0016).

### Study materials

This study used data from the OFH baseline questionnaire (version 2.2, November 2022), which all consenting participants from November 2022 were invited to complete. This questionnaire contains 286 questions that are separated into five sections: questions about “You and your household”, “Work and education”, “Lifestyle”, “Family health history”, and “Personal health history” (10).

The wording of questions in an earlier version of the questionnaire (version 1) meant that responses could not be directly compared to answers given to version 2. Participants who had completed version 1 only were removed from further analysis.

Our analysis was conducted between 28 August 2025 and 8 September 2025, using data from Data Release 11 (v11.0+ca347d7) of the OFH cohort. All data was fully anonymised prior to access for research purposes and no data that could identify individual participants could be accessed by the authors.

### Defining groups

We categorised participants who reported a history of affective disorders into three groups – bipolar disorder, depressive disorders or anxiety disorders – based on their responses to two questions in the baseline questionnaire. The first question: *“Have you ever been diagnosed with any of the following by a doctor or other health professional?”* is initially offered alongside a list of disorder categories (e.g. *Autoimmune disorder, Cancer, Neurological disorders,* etc.). Participants who selected *Mental health conditions* were then offered a second question: “*Have you ever been diagnosed with one or more of the following conditions by a professional, even if you don’t have it currently*?” alongside a list of more specific mental health conditions. We created the bipolar disorder group from those who reported a history of bipolar disorder; the depressive disorders group from participants who reported a history of depression and/or premenstrual dysphoric disorder, which we included in this group due to its predominantly affective symptomatology; and the anxiety disorders group from those who reported a history of anxiety, as well as those who reported a history of obsessive compulsive disorder and/or post-traumatic stress disorder, which were included in this group due to their association with anxiety symptoms and avoidance behaviours.

Comorbidity of different mental health problems is common (11), and the OFH questionnaire allows multiple diagnoses to be reported by the same individual. To create three mutually exclusive affective disorder groups, we applied a hierarchical approach to these groups: a diagnosis of bipolar diagnosis took precedence over the other diagnoses and a depression diagnosis took precedence over an anxiety disorder. In this way, a participant in the bipolar disorder group may have had a comorbid depressive or anxiety diagnosis, but a participant in the anxiety disorder group would not have a comorbid bipolar disorder or depressive disorder diagnosis.

We created a comparison group made up of participants who reported no past or current history of mental health problems. To do this, we removed all participants who did not answer, or who answered *Do not know, Prefer not to answer* and *Other not listed*, to the first question above as the mental health diagnosis status of these participants could not be determined. We excluded all participants who reported a history of non-affective mental health diagnoses (including eating disorder, personality disorder, psychosis, body dysmorphia, schizophrenia or schizoaffective disorder) from further analysis, along with a small proportion of participants who did not report a history of a mental health diagnosis but reported that they were taking medications for mental health conditions and/or insomnia. Participants who selected the option *None of the above* to the first question above were included in the comparison group.

### Sociodemographic, lifestyle and health-related measures

The data collected from participants in OFH provides ample opportunity to examine a wide range of sociodemographic, lifestyle and health-related measures across each group. The measures reported were selected *a priori* and include age, sex, ethnicity, educational attainment, household income, smoking status, frequency of alcohol consumption, chronotype and activity levels. Age was included as a continuous variable. The remaining measures are categorical and most comprise composite factors generated from the outputs available in the baseline questionnaire. Details on how these measures were generated and how missing data is handled for each variable can be found in the Supplementary Materials.

### Defining cardiometabolic disorders

We included the following adult-onset cardiometabolic disorders: angina, coronary artery disease, myocardial infarction, congestive heart failure, stroke, type 2 diabetes, hypertension, hypercholesterolaemia and obesity. Participants self-reported a lifetime diagnosis of any of these conditions, apart from obesity, in a similar fashion to affective disorder diagnoses. The first question: *“Have you ever been diagnosed with any of the following by a doctor or other health professional?”* is offered. Participants who select *Heart or circulatory problems* can then select *Stroke, Coronary artery/coronary heart disease, Congestive heart failure, Heart attack, Chest pain (angina), High cholesterol* and *High blood pressure (not pregnancy-related),* and those who select *Endocrine, nutritional and metabolic disorders* can select *Type 2 diabetes*. Although clinically there is likely to be overlap between several of these conditions, the OFH questionnaire provided these options in this format and so each was included as a separate outcome. It is possible for participants to select more than one of these conditions and so they should not be considered mutually exclusive. A further cardiometabolic outcome – obesity – was created from body mass index (BMI) score, which itself was calculated from self-reported height and weight at the time of questionnaire completion. Obesity was defined as a BMI score of ≥30kg/m^2^.

A cardiometabolic multimorbidity count was calculated by assigning each of the cardiometabolic conditions including obesity a ‘1’ if present and a ‘0’ if absent, and generating a total for each participant. Any individual with a score of >1 was included in the multimorbidity outcome, regardless of which specific conditions they selected. An aggregated multimorbidity outcome was generated for each of the four affective disorder groups.

### Statistical analysis

Data from OFH is accessible through the OFH Trusted Research Environment (TRE) supplied by DNAnexus, which can be accessed by authorised researchers named on specific projects approved by the OFH Access Board. The study ID for this project was OFHS240114. The responsibility for the interpretation of the information supplied by Our Future Health is the authors’ alone. The data was analysed using R v4.4.0. In line with Open Science principles, the code used to analyse the data has been uploaded [here](https://github.com/DSwiffen/OurFutureHealth-Code-Sep2025/tree/main).

Sociodemographic, lifestyle and health-related outcomes are summarised as count and proportions (N and %) for categorical variables and median and interquartile range (IQR) for continuous variables. Baseline differences between each affective disorder group and the comparison group were assessed using the Pearson’s chi-squared test with Bonferroni post hoc correction for categorical variables and one-way analysis of variance (ANOVA) with Tukey’s post hoc test for continuous variables.

Lifetime prevalence estimates (self-reported diagnoses) with 95% confidence intervals of cardiometabolic disorders were standardised by age and sex against the European Standard Population (ESP) (12). The oldest two age categories in the ESP (“85-90 years” and “90plus years”) were combined to create a single “85plus years” category as the bipolar disorder group had no participants aged over 90 years. The age range of the ESP was also narrowed to include only those aged over 18 years. Prevalence estimates were calculated for the whole sample, followed by sex disaggregated estimates.

Odds ratios with 95% confidence intervals were estimated using logistic regression comparing the odds of individual cardiometabolic disorders in each affective disorder group against those in the comparison group. Two models were created: Model 1 was adjusted for age and sex and model 2 for age, sex, ethnicity, household income, smoking status, frequency of alcohol consumption, activity levels, chronotype, and educational attainment. Age was included as a linear and quadratic term in both models. Participants with data missing in any covariate were removed prior to the logistic regression, ensuring analysis was on complete cases only. All p-values generated from logistic regression analyses were corrected for multiple comparisons using the False Discovery Rate (FDR) method to yield corrected q values.

## Results

### Sample

There were 1,781,891 participants at the time of analysis. After removing participants who had completed an earlier version of the questionnaire and those who could not be reliably categorised into any of the four groups (e.g., due to incomplete data), a total sample of 1,584,273 participants were included in the baseline comparison of sociodemographic, lifestyle and health-related factors. This comprised 8,555 participants in the bipolar disorder group, 275,764 in the depressive disorders group, 49,645 in the anxiety disorders group, and 1,250,327 in the comparison group. Age- and sex-standardised prevalence was calculated from participants with a full complement of age and sex data (n = 1,583,461). Logistic regression was completed on cases that had a full complement of data for all covariates included in the fully adjusted model (n = 1,095,492). The flow of missing data is summarised in Fig 1.

**Fig 1.**
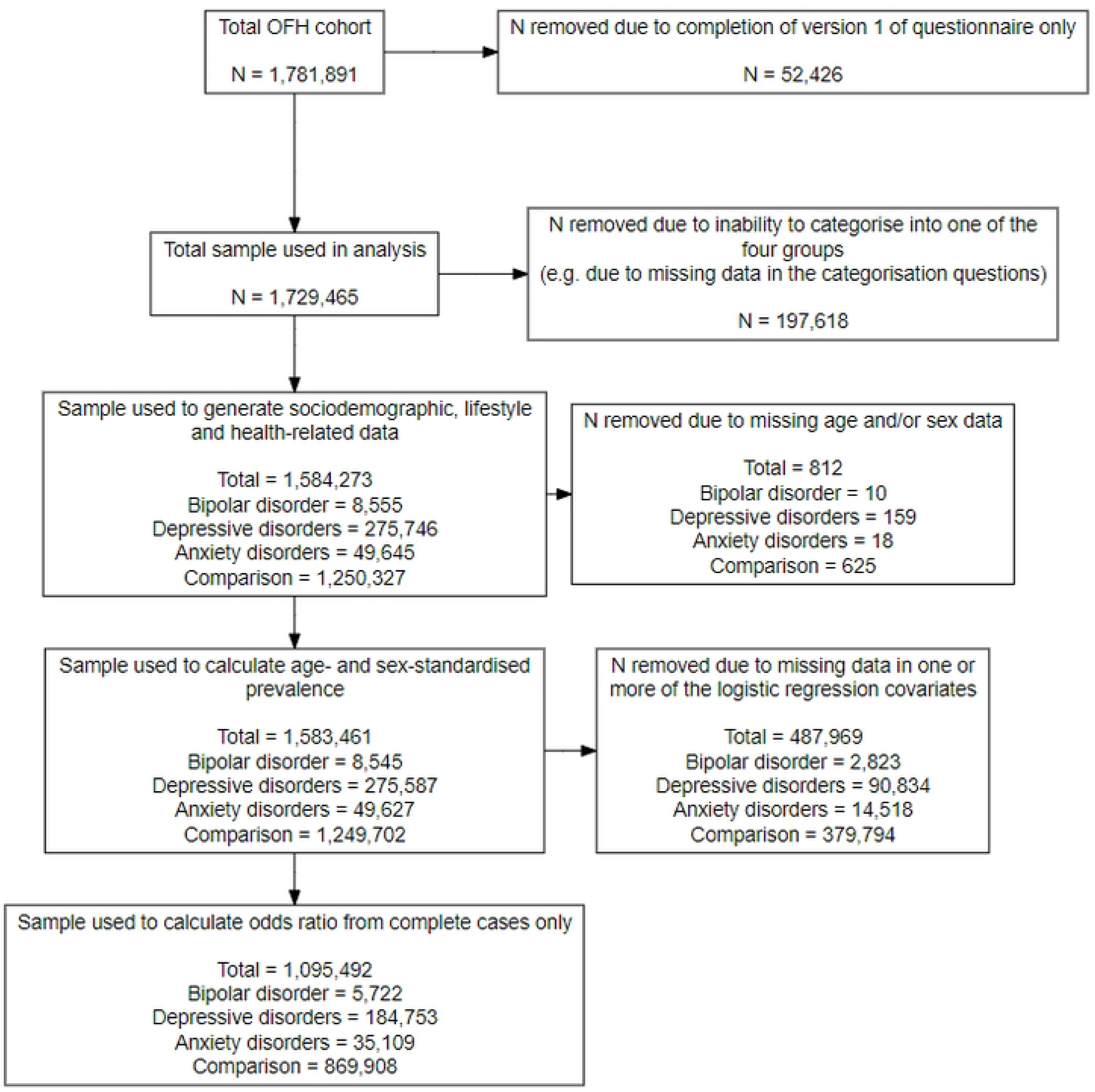
Flow of participants at each stage of analysis. A flow chart to show the sample size and missing data at each stage of analysis.

### Sociodemographic, lifestyle and health-related factors

Table 1 presents the sociodemographic, lifestyle and health-related factors across the four groups. Details on missing data per variable can be found in S1 Table.

**Table 1:** Sociodemographic, lifestyle, health-related factors by affective disorder group.

| Variable | Bipolar disorder | Depressive disorders | Anxiety disorder | Comparison |
| --- | --- | --- | --- | --- |
| **Total: *N*** | 8,555 | 275,764 | 49,645 | 1,250,327 |
| **Age: *Median (IQR)*** | 49 (36-60) | 49 (36-61) | 43 (32-57) | 57 (42-67) |
| **Female sex: *N (%)*** | 5617 (65.73) | 193120 (70.07) | 35013 (70.55) | 674987 (54.00) |
| **Ethnicity** |  |  |  |  |
| Black: *N (%)* | 102 (1.19) | 1937 (0.70) | 221 (0.45) | 21649 (1.73) |
| South Asian: *N (%)* | 174 (2.04) | 4138 (1.50) | 779 (1.57) | 48463 (3.88) |
| White: *N (%)* | 7772 (91.02) | 259096 (94.08) | 46705 (94.17) | 1114768 (89.31) |
| Mixed or multiple heritage ethnic background: *N (%)* | 248 (2.90) | 5769 (2.09) | 1012 (2.04) | 20329 (1.63) |
| Any other ethnic background: *N (%)* | 243 (2.85) | 4446 (1.61) | 880 (1.77) | 42927 (3.44) |
| **College or University degree: *N (%)*** | 4241 (50.32) | 135448 (49.68) | 29216 (59.29) | 651710 (52.83) |
| **Household income** |  |  |  |  |
| Less than £18,000: *N (%)* | 2338 (31.97) | 43092 (18.28) | 4049 (9.32) | 98849 (9.45) |
| £18,000 to £30,999: *N (%)* | 1663 (22.74) | 47640 (20.31) | 6366 (14.65) | 184049 (17.60) |
| £31,000 to £51,999: *N (%)* | 1049 (19.27) | 56885 (24.13) | 10341 (23.81) | 257275 (24.60) |
| £52,000 to £100,000: *N (%)* | 1370 (18.73) | 64702 (27.44) | 15636 (35.99) | 333923 (31.93) |
| More than £100,000: *N (%)* | 533 (7.29) | 23438 (9.94) | 7048 (16.22) | 171859 (16.43) |
| **Current or previous regular smoker (cigarettes, cigars or tobacco pipe): *N (%)*** | 5202 (62.01) | 132434 (48.91) | 18930 (38.82) | 433569 (35.39) |
| **Current alcohol drinking frequency** |  |  |  |  |
| More than once or twice per week: *N (%)* | 1674 (19.69) | 63004 (22.92) | 11352 (22.90) | 378358 (30.33) |
| Once or twice per week or less frequently: *N (%)* | 4895 (57.57) | 174780 (63.58) | 32695 (65.97) | 747876 (59.96) |
| Never: *N (%)* | 1934 (22.74) | 37109 (13.5) | 5515 (11.13) | 121155 (9.71) |
| **Evening chronotype: *N (%)*** | 4344 (57.86) | 130495 (53.30) | 21991 (48.78) | 421541 (37.62) |

| **Level of physical activity undertaken in the past 4 weeks** |  |  |  |  |
| --- | --- | --- | --- | --- |
| High level activity: *N (%)* | 781 (9.2) | 30524 (11.12) | 7944 (16.04) | 203056 (16.29) |
| Low to Medium level activity: *N (%)* | 6145 (72.4) | 210321 (76.61) | 38456 (77.67) | 978014 (78.45) |
| No physical activity: *N (%)* | 1561 (18.39) | 33680 (12.27) | 3111 (6.28) | 65596 (5.26) |

Each affective disorder group was compared with the comparison group to generate p-values. For each variable these were $p < 0.0001$ . Continuous variables (Age) were compared using one-way analysis of variance (ANOVA) with Tukey's post hoc test whilst categorical variables (Female sex, Ethnicity, College or University degree, Household income, Current or previous regular smoker, Current alcohol drinking frequency, Evening chronotype and Level of physical activity taken in the past 4 weeks) were compared using Pearson's chi-squared test with Bonferroni post hoc correction. Details on the derivations of each variable are provided in Supplementary Materials.
Missing data counts and proportions are provided in S1 Table.

All baseline variables for each affective disorder group were significantly different from the comparison group (p <0.001). Compared to those without any lifetime history of mental disorder, participants with affective disorders tended to be younger, more often female, more often of White ethnicity, more likely to have ever smoked, less frequent consumers of alcohol, more often of evening chronotype and less physically active.

### Prevalence of cardiometabolic disorders

The standardised prevalence [95% confidence interval] of any cardiometabolic disorder was higher in each of the affective disorder groups than in the comparison group. In Table 2, we report that for bipolar disorder this was 41.14% [39.36 42.93], for depressive disorders 36.75% [36.49, 37.02], for anxiety disorders 28.79% [28.38, 29.38] and for the comparison group 24.75% [24.66, 24.83].

**Table 2:** Standardised prevalence of cardiometabolic disorder by affective disorder group.

| **Cardiometabolic disorder\*** | **Bipolar disorder: % (95% CI)** | **Depressive disorders: % (95% CI)** | **Anxiety disorders: % (95% CI)** | **Comparison: % (95% CI)** |
| --- | --- | --- | --- | --- |
| **Any** | 41.14 (39.36, 42.93) | 36.75 (36.49, 37.02) | 28.79 (28.38, 29.38) | 24.75 (24.66, 24.83) |
| **Angina** | 3.32 (2.91, 3.72) | 2.33 (2.21, 2.44) | 1.39 (1.16, 1.62) | 0.98 (0.96, 1.00) |
| **Coronary artery disease** | 2.29 (1.78, 2.80) | 1.87 (1.77, 1.98) | 1.44 (1.22, 1.65) | 1.17 (1.15, 1.19) |
| **Myocardial infarction** | 1.67 (1.34, 2.00) | 1.40 (1.30, 1.49) | 0.96 (0.79, 1.14) | 0.92 (0.90, 0.93) |
| **Congestive heart failure** | 0.71 (0.49, 0.94) | 0.41 (0.36, 0.46) | 0.31 (0.18, 0.44) | 0.20 (0.19, 0.21) |
| **Stroke** | 1.58 (1.30, 1.87) | 1.39 (1.31, 1.48) | 1.01 (0.81, 1.22) | 0.83 (0.81, 0.85) |
| **Hypertension** | 14.24 (13.33, 15.15) | 14.19 (13.99, 14.39) | 11.68 (11.21, 12.15) | 9.21 (9.16, 9.26) |
| **Hypercholesterolemia** | 10.46 (9.63, 11.29) | 9.70 (9.53, 9.88) | 7.75 (7.34, 8.16) | 5.73 (5.69, 5.77) |
| **Type II diabetes** | 5.69 (5.16, 6.22) | 3.80 (3.69, 3.91) | 1.59 (1.48, 1.71) | 1.78 (1.76, 1.81) |
| **Obesity (BMI >30)** | 29.57 (27.88, 31.25) | 24.73 (24.50, 24.95) | 18.14 (17.71, 18.58) | 15.48 (15.41, 15.55) |
| **Cardiometabolic multimorbidity** | 12.50 (11.63, 13.37) | 11.80 (11.62, 11.99) | 9.18 (8.75, 9.61) | 6.89 (6.85, 6.94) |

Prevalence estimates are standardised by age and sex against the European Standard Population
\*Cardiometabolic disorders are not mutually exclusive

Standardised prevalence of cardiometabolic disorders in the four groups are shown in Table 2 and Fig 2. All cardiometabolic disorders were more prevalent in the bipolar disorder and depressive disorders groups than in the comparison group. Most cardiometabolic disorders also had higher prevalence in the anxiety disorders group than in the comparison group, except type 2 diabetes which had a marginally lower prevalence in those with anxiety disorders, and myocardial infarction, congestive heart failure and stroke, which were similarly prevalent in the anxiety disorders group as in the comparison group.

**Fig 2.**
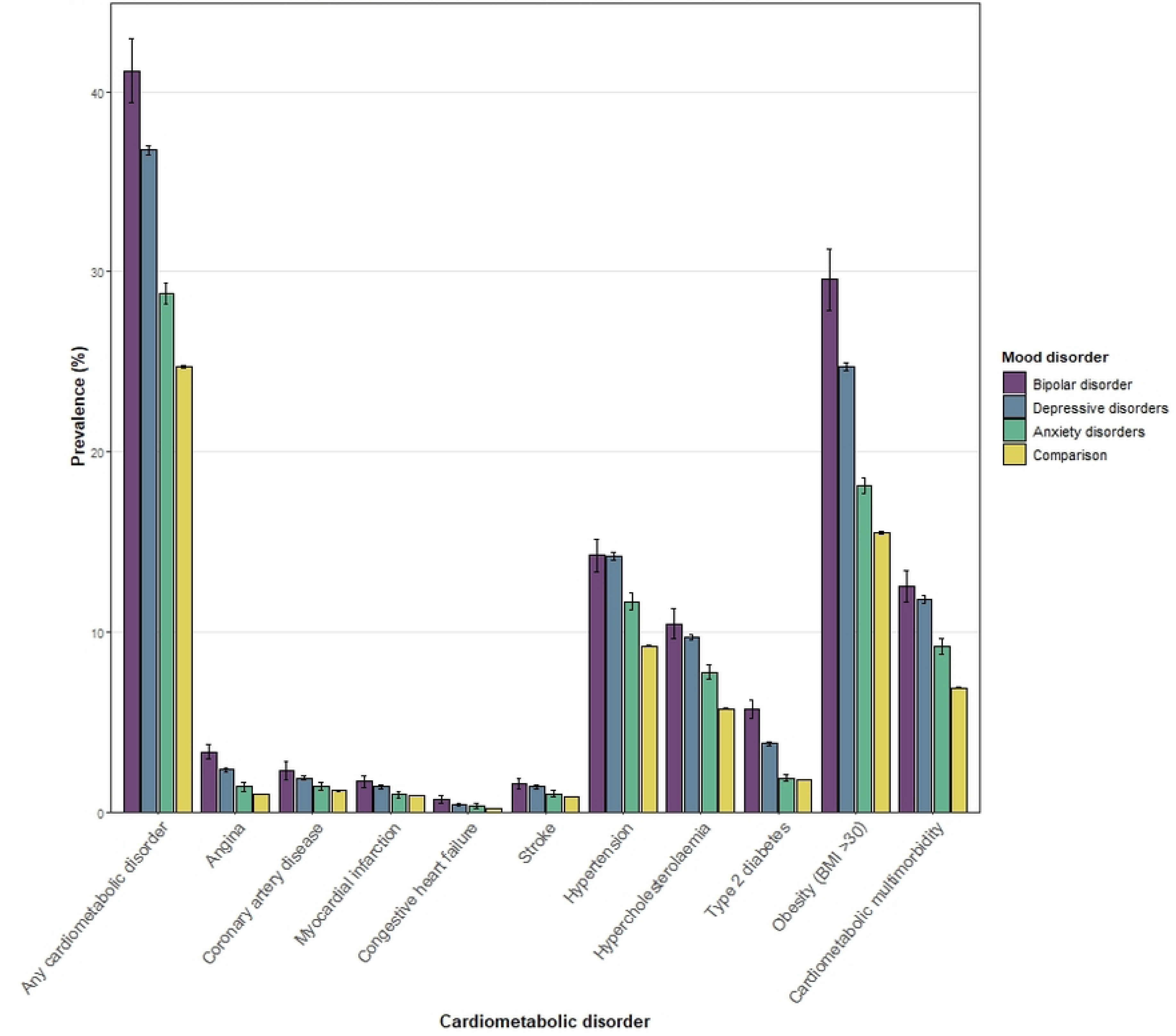
Standardised prevalence of cardiometabolic disorders by mood disorder. A bar chart to show age- and sex-standardised prevalence with 95% confidence intervals of cardiometabolic disorders by affective disorder and comparison groups.

Sex disaggregated standardised prevalence of cardiometabolic disorders is presented in Fig 3. Across all affective disorders and the comparison group, males reported higher prevalence of all cardiometabolic disorders than females, with the exception of obesity.

**Fig 3.**
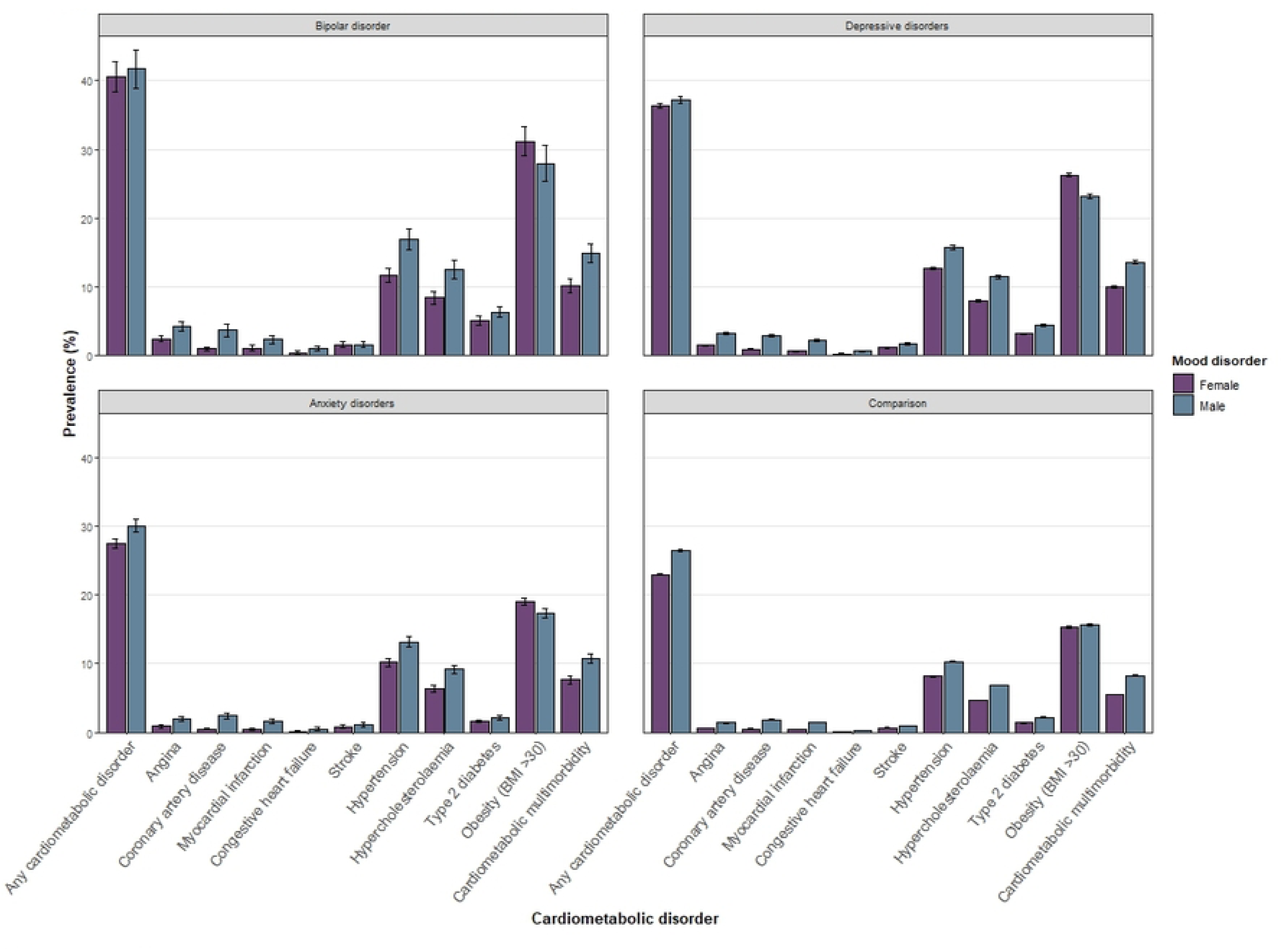
Standardised prevalence of cardiometabolic disorders by mood disorder and sex. A bar chart to show age- and sex-standardised prevalence with 95% confidence intervals of cardiometabolic disorders separated by sex for each affective disorder group and the comparison group.

### Logistic regression

Fig 4 and S2 Table show the adjusted odds ratios with 95% confidence intervals for individual cardiometabolic disorders in each affective disorder group compared to the comparison group. FDR- corrected q-values for these comparisons are also provided in S2 Table. In general, the odds of cardiometabolic disorders were significantly greater in the affective disorder groups, and were particularly high in the bipolar disorder group, even after adjusting for a range of covariates.

**Fig 4.**
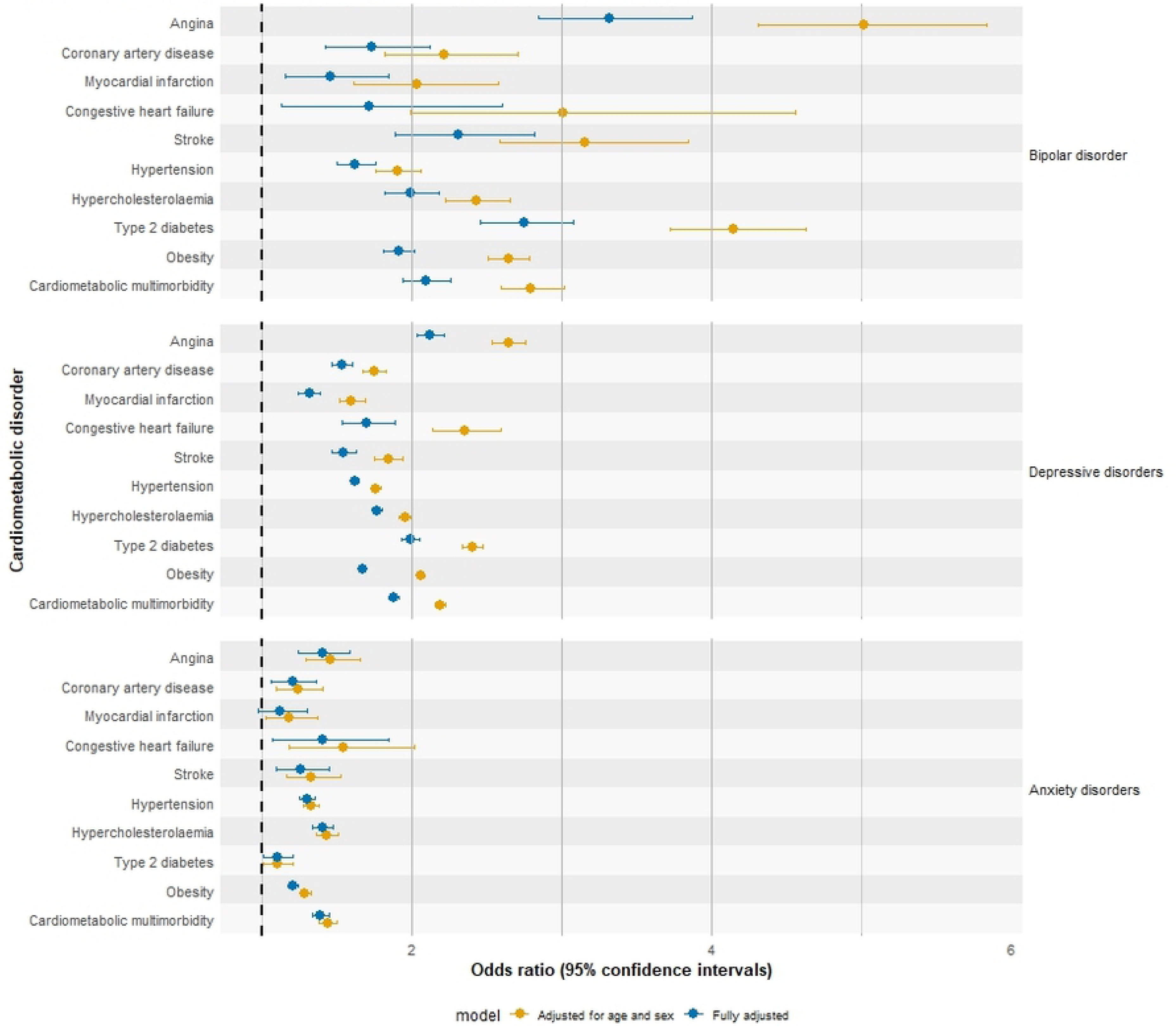
Odds ratios of cardiometabolic disorder by affective disorder group. Forest Plot to show the adjusted odds ratios of cardiometabolic disorder diagnoses in each affective disorder group (versus the comparison group).

The first model is adjusted for age and sex only. The fully adjusted model is adjusted for age, sex, ethnicity, household income, smoking status, frequency of alcohol consumption, activity levels, chronotype, and educational attainment.

Both models contain complete cases only. Participants with data missing for any of the covariates were removed prior to analysis. S3 Table contains a comparison of baseline characteristics for complete cases vs incomplete cases. *N* for both models = 1,095,492.

Based on the fully adjusted model, people with bipolar disorder generally had the highest odds of each cardiometabolic disorder, followed sequentially by people with depression and people with anxiety, compared to the comparison group. This is reflected in the odds ratios for cardiometabolic comorbidity, which are highest in bipolar disorder (adjusted odds ratio 2.10 [95% confidence interval 1.94, 2.26, q <0.001]), followed by depression (1.88 [1.85, 1.91, q <0.001]) and then anxiety (1.39 [1.34, 1.45, q <0.001]).

In people with bipolar disorder, the highest increased odds are seen for angina (3.32 [2.85, 3.87, q <0.001]), type 2 diabetes (2.75 [2.46, 3.08 q <0.001]) and stroke (2.31 [1.89, 2.82, q <0.001]). In people with depression, the highest increased odds are also seen for angina (2.13 [2.03, 2.22, q <0.001]) and type 2 diabetes (1.99 [1.93, 2.05, q <0.001]). For people with anxiety, angina (1.40 [1.24, 1.59, q <0.001]), hypercholesterolaemia (1.41 [1.34, 1.48, q <0.001]) and congestive heart failure (1.41 [1.07, 1.85, q = 0.01]) had the highest increased odds, while the association with type 2 diabetes is smaller ((1.11 [1.02, 1.21, q = 0.02]).

## Discussion

This is the first report within the landmark Our Future Health cohort of the prevalence of cardiometabolic disorders in individuals with a lifetime self-reported diagnosis of bipolar disorder, depression or anxiety. The highest prevalence of cardiometabolic disorders was observed in those with bipolar disorder. Males were observed to have the highest prevalence of all cardiometabolic disorders apart from obesity. In models adjusted for a range of lifestyle and sociodemographic factors, we identified higher odds of cardiometabolic disorders in all three affective disorder groups. The strongest associations were found for those with bipolar disorder, followed by those with depressive disorders, and those with anxiety disorders showed the smallest associations. The cardiometabolic disorder most strongly associated with affective disorder was angina, with type 2 diabetes also showing a strong association with bipolar disorder and depression and weak association with anxiety disorders.

Our analysis was conducted on one of the largest health cohorts in the world, providing a high degree of statistical power. Large cohort studies have the risk of self-selection bias and previously have been found to have samples that tend to skew older and healthier (13). Our Future Health, however, has implemented strategies to ensure nationally-representative recruitment that is more reflective of UK census data. Whilst these efforts will make findings more generalisable to the UK population, the current recruited cohort is made up of relatively more females, fewer younger people and more people aged 50-79 years than the UK population (14). As with all cohort studies, issues around representative recruitment and selection bias will still be relevant.

The findings from our analysis are largely consistent with previous work, however many of the associations found are of greater magnitude than previously reported. Evidence from a recent umbrella review and meta-analysis found associations between bipolar disorder and hypertension (equivalent odds ratio, eOR [95% confidence interval] = 1.28 [1.02, 1.60]), obesity (eOR 1.64 [1.30, 1.99]) and type 2 diabetes (eOR 1.98 [1.55, 2.52]) (15) that were all of lower magnitude than our findings (the odds of type 2 diabetes, for example, was almost three times greater in our bipolar disorder group versus comparison). For depression and anxiety, our findings are again largely comparable with previous reports. For example, increased relative risk for coronary artery disease (RR 1.41 [1.23, 1.61]) and congestive heart failure (RR 1.35 [1.11, 1.64]) in anxiety disorder has been noted previously (16) and a previous meta-analysis also found similarly increased odds of cerebrovascular disease in depression (OR 1.64 [0.96, 2.78]) (17).

There are many converging aetiological mechanisms thought to link affective and cardiometabolic disorders. For example, chronic hypothalamic-pituitary-adrenal (HPA) axis dysfunction and hypercortisolaemia, as seen in anxiety, depression and bipolar disorder, may contribute to insulin resistance and the risk of diabetes (6, 18, 19), and prolonged stress, as is characteristic of depression or mania in bipolar disorder, can lead to autonomic nervous system dysfunction, impairing cardiac contractility, heart rate variability and regulation of blood pressure (6). The relationship is bidirectional, as acutely elevated cortisol levels and increased inflammation likely contribute to the high rates of depression seen after myocardial infarction and stroke (20, 21). The circadian clock, commonly disrupted in affective disorders (22), may also play an important part as it influences adipose tissue metabolism, HPA axis function, and blood pressure and heart rate regulation, all of which can impact cardiometabolic health (23, 24). Although we corrected for chronotype in our analysis, other circadian disruptions such as shift work, late time eating and light exposure at night can impact circadian function and may in turn increase the risk of cardiometabolic and affective disorders. This complex and overlapping pathophysiology of affective and cardiometabolic disorders highlights the need for integrated approaches in assessment and management, as well as further research into their shared mechanisms to underpin development of future treatments.

### Strengths and limitations

We adjusted our analyses for a range of relevant confounders, including age, sex, smoking, activity levels, and measures of socioeconomic status, but this list was not exhaustive. Predictors that are known to impact metabolic and cardiovascular health that we did not account for include dietary intake and psychotropic medication use (25, 26). Generally, antipsychotics and mood stabilisers – which are used more frequently in bipolar disorder – have a greater effect on weight gain, dyslipidaemia and glucose dysregulation than antidepressants (27). It is therefore possible that the psychotropic medications may influence cardiometabolic outcomes of the bipolar group more than in the depressive and anxiety disorders groups. Evidence from people with newly diagnosed bipolar disorder, however, highlights that the connection between cardiometabolic disorders and affective disorders cannot be solely explained by exposure to medications (28).

Our classification of affective disorder groups and cardiometabolic disorders may differ from that of other studies in this area. Whilst there are pragmatic benefits to using self-reported lifetime diagnosis over clinician-rated scales for this purpose (such as cost and time considerations), there may be self-report biases. Previous studies have demonstrated moderate agreement at best between symptom-based and self-reported diagnostic measures for anxiety and depression in previous cohort studies (29) highlighting the risk that self-reported diagnosis may result in some participant misclassification. In addition, we grouped affective disorders into three relatively broad diagnostic groups, which may have affected our findings. Previous evidence demonstrates significantly greater cardiovascular and cerebrovascular mortality in bipolar disorder type I compared to bipolar disorder type II (30), indicating the potential for important differences within each mood disorder group. The grouping of affective disorders into the three groups has pragmatic benefits for analysis and interpretation, but risks missing more subtle differences that might exist. Such differences may be useful to explore in more granular future analyses.

An important limitation of our study is the number of participants that were excluded from logistic regression due to missing data in one or more of the included covariates. Household income and chronotype had the most missing data, mainly due to participants choosing the options *Prefer not to answer* or *Do not know* for these questions. A comparison of the baseline characteristics between complete cases that were included in logistic regression and incomplete cases indicated that the complete cases are more educated, tend to have higher household incomes, are less likely to have an evening chronotype and are more likely to have engaged in a high-level physical activity in the past four weeks. They are, however, slightly older and are more frequent drinkers of alcohol. The rates of a regular smoking history were very similar between complete and incomplete cases. These differences highlight that the sample used in the logistic regression may not be representative of the UK population as a whole.

A final limitation of our study is that we are not able to determine causality from cross-sectional data. Although we identified associations between affective and cardiometabolic disorders, it is not possible to comment on directionality. Further prospective analyses and causal inference approaches (such as Mendelian randomisation) are needed to investigate causal mechanisms and to explore the direction of the association between affective and cardiometabolic disorders.

There are several possibilities for future work in this area, including (but not limited to) a more detailed analysis of sex differences, analysis of the importance of modifiable risk factors such as sleep, activity, social connection and diet, and the importance of medication effects (both psychotropic medication and other medication classes).

## Conclusion

We have identified associations between three broadly defined self-reported groups of affective disorders (bipolar disorder, depressive disorders and anxiety disorders) and increased risk of multiple cardiometabolic disorders. Differences between the groups remained after adjusting for relevant sociodemographic, lifestyle and health-related factors but future studies are required to explore causality and mechanisms. Our analyses also highlighted a particularly high burden of cardiometabolic comorbidity for bipolar disorder and especially in men, which warrants further systematic investigation.

## Data Availability

Data associated with this research are not publicly available. Access to Our Future Health data is restricted to registered researchers who make a successful study application to the Access Board for the cohort. The authors do not have permission to share this data.

https://github.com/DSwiffen/OurFutureHealth-Code-Sep2025.git

## Acknowledgements

This study makes use of de-identified data held by Our Future Health. We would like to acknowledge all the research participants who have donated their data to the Our Future Health research programme.

## Author contributions

DS, AMRL, AR, CS, KFMM and DJS were involved in the conceptualisation of the project. Methodology was devised by DS, KF and DJS. Data curation and formal analysis was completed by DS, with support from AMRL, AR, CS and KF. Preparation of the original draft was completed by DS. Review and editing of the original and subsequent drafts were completed by AMRL, AR, CS, KRMM, KF and DJS. Supervision was provided by DJS.

## Supporting information

**S1 Table. Sociodemographic, lifestyle, health-related factors and missing data per variable by affective disorder group.** Sociodemographic, lifestyle and health-related factors are compared across each affective disorder groups and comparison group, with frequency and proportion of data classed as missing per covariable included.

**S2 Table. Odds ratio (with 95% confidence intervals and corrected q-values) for cardiometabolic disorders by mood disorder group versus comparison group.** The odds ratio for models 1 and 2 of the logistic regression analysis are presented. Model 1 odds ratio are adjusted for age and sex. Model 2 odds ratio are adjusted for age, sex, ethnicity, household income, smoking status, frequency of alcohol consumption, activity levels, chronotype and educational attainment. *N* for both models = 1,095,492. †p-values have been corrected using the False Discovery Rate method for multiple statistical tests to generate corrected q-value.

**S3 Table. Baseline characteristics of participants with complete data versus participants with incomplete data**. Baseline sociodemographic, lifestyle and health-related factors of participants who have a full complement of data for each included covariate are compared against those who had data missing for at least one covariate. For each covariate under “Participants with incomplete data”, the percentage reported is based on participants who did not have missing data for that covariate.

**S1 File. Defining outcome measures for sociodemographic, lifestyle and health-related variables.** Further details are provided on how the covariates included in the logistic regression were coded based on outputs from the Our Future Health baseline questionnaire.

